# Colorectal cancer in patients with single versus double positive faecal immunochemical test results: A retrospective cohort study

**DOI:** 10.1101/2020.05.11.20097881

**Authors:** Tian Zhi Lim, Jerrald Lau, Gretel Jianlin Wong, Ker-Kan Tan

## Abstract

**BACKGROUND:** Screening for colorectal cancer (CRC) using the faecal immunochemical test (FIT) is widely advocated. Few studies have compared the rate of detecting colonoscopic pathologies in single compared to double FIT-positive follow-up colonoscopy-compliant individuals in a two-sample national FIT screening program.

**OBJECTIVE:** To compare CRC incidence in double FIT-positive versus single FIT-positive individuals using a retrospective cohort of patients from a tertiary hospital in Singapore.

**DESIGN:** Retrospective cohort study.

**SETTING:** Data was extracted from one regional acute hospital in Singapore.

**PARTICIPANTS:** 1,539 FIT-positive individuals from the national FIT screening program who were referred to the hospital from 1st January 2017 to 31st September 2019 for follow-up consultation and diagnostic colonoscopy.

**MEASUREMENTS:** The exposure of interest was a positive result on both FIT kits. The main outcome was a follow-up diagnostic colonoscopy finding of CRC. The secondary outcome was a diagnostic colonoscopy finding of a colorectal polyp.

**RESULTS:** Incidence density of CRC was 1.53 (95% CI = 0.61, 3.15) and 17.88 (95% CI = 11.67, 26.19) per 100,000 person-months, in the single and double FIT-positive group, respectively. This resulted in an incidence rate ratio of 11.71 (95% CI = 5.25, 29.08). Colorectal polyp detection was significantly higher (p < 0.01) in the double (107 of 157 participants; 68.2%) compared to the single (310 of 585 participants; 53.0%) FIT-positive group.

**LIMITATIONS:** The key limitation of this study was the relatively small cohort derived from a single regional hospital, as this had the effect of limiting the number of incident cases, resulting in comparatively imprecise CIs.

**CONCLUSIONS:** Double FIT-positive individuals are significantly more likely to have a colonoscopy finding of incident CRC or premalignant polyp than single FIT-positive individuals. Clinicians and policymakers should consider updating their CRC screening protocols accordingly.

**FUNDING SOURCE:** This study was supported by the Singapore Population Health Improvement Centre (SPHERiC) [NMRC/CG/C026/2017_NUHS]. The funders had no role in the study design, execution, analyses, interpretation of the data, or decision to submit results. We confirm that all authors had full access to all of the data (including statistical reports and tables) in the study and can take responsibility for the integrity and accuracy of the data analysis.

## INTRODUCTION

Colorectal cancer (CRC) screening has been shown to improve oncological outcomes by earlier detection and treatment of premalignant adenomatous polyps or CRC itself (1). CRC screening modalities include the faecal immunochemical test (FIT), colonoscopy, flexible sigmoidoscopy, computed tomographic colonography and stool DNA testing. However, FIT remains the most cost-effective (1). In the event that any FIT turns positive, a follow-up colonoscopy is recommended to complete the colonic evaluation (1, 2).

Singapore has adopted two-sample FIT in its national CRC screening program as this has been shown to increase sensitivity by potentially detecting neoplasms that may have been missed in one-sample FIT (3, 4). The national CRC screening program advises age-eligible Singapore residents to complete a free two-sample FIT kit screening every year (3). Should either sample be positive, the individual would be contacted by a dedicated hospital coordinator to arrange for consultation and colonoscopy with a colorectal surgeon or gastroenterologist (5).

Individuals with two positive FIT samples (“double FIT-positive”) – which would have been sampled over two days – should intuitively be more likely to have colonic pathologies compared to individuals with only one positive FIT sample (“single FIT-positive”). However, few studies have examined these outcomes (4, 6). Thus, this study aimed to compare CRC incidence in double FIT-positive versus single FIT-positive individuals using a retrospective cohort of patients from a tertiary hospital in Singapore.

## METHODS

### Ethical approval

This study was ethically approved by the National Healthcare Group’s Domain Specific Review Board (NHG DSRB; Reference number: 2017/01260) in accordance with the Declaration of Helsinki.

### Patient and public involvement statement

This research was done without patient involvement. Patients were not invited to comment on the study design and were not consulted to develop patient relevant outcomes or interpret the results. Patients were not invited to contribute to the writing or editing of this document for readability or accuracy.

### Study design and participants

The study population (N = 1,641) was a retrospective cohort comprising all individuals who (I) had completed and submitted two FIT kits under the national CRC screening program, (II) received at least one positive FIT-result, and (III) were referred to the National University Hospital (NUH) between 1^st^ January 2017 to 31^st^ September 2019 for follow-up consultation and diagnostic colonoscopy. The index date was the date of referral to the hospital.

Individuals were excluded from the study if they (I) had missing or incomplete results for either FIT kit, (II) had transferred to another hospital, or (III) had not yet completed the scheduled follow-up colonoscopy at the point of data extraction (12th December 2019). This resulted in a final sample of 1,539 participants, or 6.2% loss to follow up.

### Primary exposure and outcomes of interest

The exposure of interest was defined as a positive result on both FIT kits. The main outcome (incident CRC cases) was defined as a follow-up diagnostic colonoscopy finding of a CRC, regardless of staging. The secondary outcome was defined as a follow-up diagnostic colonoscopy finding of a colorectal polyp. Colonoscopy reports were verified by using the hospital’s electronic medical records system

### Other participant factors

Apart from the main exposure and outcome of interest, our data included participants’ ethnicity (defined as Chinese, Malay, Indian, or Others), age, date of two-sample FIT kit collection, date of referral to NUH, and date of follow-up medical consultation and colonoscopy.

### Statistical analysis

For demographics, frequencies and proportions were used to report ethnicity and compliance to follow-up colonoscopy. Median and interquartile range were used to summarise participant age. For colonoscopy-compliant participants, median and interquartile range was used to report follow-up time (in days) between date of referral to the hospital and date of colonoscopy. Incident CRC cases were summarised using frequencies and proportions.

To examine possible effects of confounding, chi-square tests were used to compare ethnicity and colonoscopy compliance, and Mann-Whitney U tests for age and follow-up time, between single and double FIT-positive groups.

A chi-square test was used to compare incident CRC cases and polyp detection between the groups. Incidence density (ID) of CRC cases was derived using person-months at risk, calculated from month of birth to the month of colonoscopy. Incidence rate ratio (IRR) was used to compare relative risk of CRC between the single FIT-positive and double FIT positive groups. Confidence intervals (95% CIs) were reported for both ID and IRR.

## RESULTS

The median age of the sample was 65 years (IQR = 58 – 71 years). Overall compliance to follow-up colonoscopy was 50.4%. The distribution of sample demographics, colonoscopy compliance, and outcomes of interest between single and double FIT-positive patients can be found in Table 1.

**Table 1.** Distribution of age, ethnicity and follow-up colonoscopy compliance between single FIT-positive and double FIT-positive participants

|  | Single FIT-positive<br>(*n* = 1184) | Double FIT-positive<br>(*n* = 355) | *p*-value |
| --- | --- | --- | --- |
| Median age, years<br>(IQR) | 64.0<br>(57.2 – 70.2) | 68.0<br>(60.5 – 73.0) | < 0.01 |
| Ethnicity, *n* (%) |  |  | 0.91 |
| Chinese | 1090<br>(92.1) | 324<br>(91.3) |  |
| Malay | 50<br>(4.2) | 18<br>(5.1) |  |
| Indian | 29<br>(2.4) | 8<br>(2.3) |  |
| Other | 15<br>(1.3) | 5<br>(1.4) |  |
| Colonoscopy<br>compliance, *n* (%) | 592<br>(50.0) | 183<br>(51.6) | 0.61 |
| Follow-up time<br>from referral to<br>colonoscopy,<br>median days (IQR) | 49<br>(29 – 75) | 43<br>(26 – 73) | 0.12 |
| Incident CRC<br>cases, *n* (%)* | 7<br>(1.4) | 26<br>(14.2) | < 0.01 |
| Colonoscopy with<br>polyp finding, *n*<br>(%)** | 310<br>(53.0) | 107<br>(68.2) | < 0.01 |

\*Note: Denominator for proportions were based on colonoscopy-compliant participants only.
\*\*Note: Denominator for proportions were based on colonoscopy-compliant participants, and excludes incident CRC cases detected.

Median follow-up time between date of referral to the hospital and date of colonoscopy was calculated for colonoscopy-compliant patients between the single (*n* = 592; median = 49 days, IQR = 29 – 75 days) and double FIT-positive (*n* = 183; median = 43 days, IQR = 26 – 73 days) groups; *U* = 50,065.0, *p* = 0.12.

Incidence density of CRC was 1.53 per 100,000 person-months (95% CI = 0.61, 3.15) in the single- and 17.88 per 100,000 person-months (95% CI = 11.67, 26.19) in the double FIT positive group. This resulted in an incidence rate ratio of 11.71 (95% CI = 5.25, 29.08) (refer to Table 2).

**Table 2.** Incidence density of colorectal cancer, and incidence rate ratio, between single FITpositive and double FIT-positive colonoscopy-compliant participants

|  | Incident cases | Person-months at risk | ID per 100,000 person-months (95% CI) | IRR (95% CI) |
| --- | --- | --- | --- | --- |
| Single FIT-positive<br>(*n* = 592) | 7 | 458507 | 1.53<br>(0.61, 3.15) |  |
| Double FIT-positive<br>(*n* = 183) | 26 | 145446 | 17.88<br>(11.67, 26.19) | 11.71<br>(5.25, 29.08) |

## DISCUSSION

In colonoscopy-compliant patients, the double FIT-positive group was approximately 12 times more likely to have CRC and a significantly higher incidence of colorectal polyps than the single FIT-positive group. These findings have significant implications to several stakeholders.

There is the need to expedite colonoscopy for those who are double FIT-positive to ensure prompt detection of CRC and premalignant polyps. With increasing advocacy on CRC screening and being current to screening recommendations, a higher number of patients who are at least single FIT-positive is expected if screening uptake increases. This could put strains on healthcare systems with finite facilities for colonoscopy, resulting in a need to prioritise which patients should more urgently undergo the procedure.

At the national level, countries that are still advocating CRC screening using only one-sample FIT should reconsider their approach. Double FIT-positive is likely to have increased specificity, resulting in fewer false positive findings during follow-up (4, 6). Recent findings have also highlighted the significantly higher positive predictive value of double FIT-positive compared to single FIT-positive in detecting CRC (4). Although compliance rates of individuals performing two-sample FIT over two separate days are likely to be slightly lower than one-sample FIT, this can be managed through public education and targeted interventions (6). Countries concerned about costs to the health system should consider performing a cost-effectiveness analysis, which will likely favour a two-sample FIT program (6).

More work is also required to reinforce the importance of prompt follow-up after a positive FIT result. As observed in our study and the literature, colonoscopy compliance rate after a positive FIT is approximately 50% (7). Using simple arithmetic, another 33 CRC cases could have been diagnosed and managed promptly should there have been perfect compliance.

Prior literature has suggested a combination of patient and provider factors influencing follow-up compliance (7). As these have been known to vary between populations, public health professionals and policymakers must understand population-specific barriers and facilitators in order to develop tailored, cost-effective interventions.

The key strengths of this study were (I) the representativeness of the sample to the average risk study population recommended to our hospital via the national screening programme, with a loss to follow up under 10%, and (II) the ability to verify outcomes of interest via the hospital’s clinical database. The key limitation of this study was the relatively small cohort derived from a single regional hospital, as this had the effect of limiting the number of incident cases, resulting in comparatively imprecise CIs. Our findings should nonetheless be representative of national demographics as FIT-positive hospital referrals are performed geographically, and population sociodemographic characteristics should be similarly distributed across Singapore (8). Although there is certainly the need to validate our findings in larger two-sample FIT cohorts, the significant difference in the rates of CRC detected should prompt clinicians and policymakers to consider examining this issue with greater urgency.

## Data Availability

The data that support the findings of this study are available from the corresponding author, KKT, upon reasonable request.

